# Identifying risk factors for Lassa fever infection in Sierra Leone, 2019-2021

**DOI:** 10.1101/2024.04.11.24305423

**Authors:** Daniel Juma Sama, Najmul Haider, Javier Guitian, Abdinasir Yusuf Osman, Francine Ntoumi, Alimuddin Zumla, Richard Kock, Rashid Ansumana

## Abstract

**Background:** Lassa fever (LF) virus (LASV) is endemic in Sierra Leone and poses a significant public health threat to the region; however, no risk factors for LASV infection have been reported in Sierra Leone. The objective of this study was to identify the risk factors for LASV infection in an endemic community in Sierra Leone.

**Methods:** We conducted a case-control study by enrolling 37 laboratory-confirmed LF cases identified through the national LF surveillance system in Sierra Leone, and 140 controls resided within a one-kilometre radius of the case household. We performed conditional multiple logistic regression analysis to identify the risk factors for LASV infection.

**Results:** Of the 37 cases enrolled, 23 died (62% case fatality rate). Cases were younger than controls (19.5 years vs 28.9 years, p<0.05) and more frequently female (64.8% vs. 52.8%). Compared to the controls, LF cases had contact with rodents (rats or mice) in their households more frequently in the preceding three weeks (83.8% vs.47.8%). Households with a cat reported a lower presence of rodents (73% vs 38%, p<0.01) and contributed to a lower rate of LF (48.6% vs 55.7%) although not statistically significant (p=0.56). The presence of rodents in the households (Matched Adjusted Odds Ratio [mAOR]: 11.1), and younger age (mAOR: 0.99) were independently associated with LASV infection.

**Conclusion:** Rodent access to households is likely a key risk factor for LASV infection in rural Sierra Leone and potentially in other countries within the West African region. Controlling rodent access to households might help reduce household-level LASV infection in Sierra Leone.

## Introduction

Lassa fever (LF) virus (LASV) is a viral zoonotic illness caused by an arenavirus and is responsible for severe haemorrhagic fever characterized by fever, muscle aches, vomiting, chest, and abdominal pain with several complications including deafness [1]. The disease is endemic in West Africa including Sierra Leone (SL) [2–6]. In a 1980s estimate, LF was reported to infect approximately 200,000-300,000 people and cause 5,000-10,000 human deaths each year in West Africa [7]. However, in the last four decades, the population in Sub-Saharan Africa (SSA) has doubled and crop production has intensified resulting in losses of forest areas and destruction of ecosystems, which could have created conditions more favourable for LASV infection. A 2020 model estimated an annual incidence of more than 800,000 LF cases in West Africa [8].

*Mastomys natalensis* is the primary reservoir of LASV [9,10], however, two other species, *Mastomys erytholeucus,* and *Hylomyscus pamfi* were recently identified as a reservoir of LASV [11,12]. Programs on rodent control to fight against LASV conducted in West Africa listed several drawbacks in the successful elimination of rodents [13] including, the prolificacy of *M. natalensis* with a mean litter size of 9.2 (range: 3-14), the ability of some rodents to survive with a lethal dose of baited poison, lack of implantation of recommendations by communities (for whatever reason), availability of alternate food that helps rodents to escape baited food, the porosity of the houses/rooms allowing the rodent to enter and live, and low number of natural predators of rodent in the community. In Sierra Leone, most of the towns and villages are embedded in a fragmenting forest or bush environment-creating an opportunity for invasion of species able to adapt to human conditions and housing. Most dwelling houses in SL store primary crops and their residues from subsistence agriculture provide an easy food source to increase increasing the likelihood of human contact with rodents and their faeces or urine.

Humans are believed to get infections through touching objects contaminated with rodent urine, breathing aerosolized particles, being bitten by rodents, or, consuming rodents [14–16]. Human-to-human transmission can occur occasionally in hospital settings and the community [17–19]. Earlier studies identified several risk factors mostly associated with human-human transmission [20]. Kerneis et al (2009) reported living with someone with a haemorrhagic and receiving an injection in past years as a risk factor for LASV infection [20]. Another study from Nigeria reported that the LF cases had a history of consuming rodent-contaminated food (56%) or being exposed to LF-infected individuals (15.8%) [21].

Risk factors related to human-to-human infection further mean that the enrolled cases were not index case. Furthermore, most of the risk factors identified were reported through a cross-sectional study thus raising the ambiguity of temporality of the cases and exposure. Nonetheless, no risk factors are reported in Sierra Leone. Thus, the objective of this study was to estimate the risk factors for LASV infection in an endemic district of Sierra Leone to synthesize evidence to support policies and programs to prevent household-level exposure of LASV to humans.

## Methods

We collected the list of Lassa fever cases identified between January 2019 and December 2021 from the National Lassa fever surveillance unit based in Kenema Hospital, Sierra Leone. Our team consists of a research officer and a research assistant. Both received training on the administration of the questionnaire. The questionnaire was pre-tested in a similar village in the Kenema district and modified based on the field observation. We defined a case as a person who has been confirmed with presented positive results for LASV detection by either RT-PCR or serology (IgM or IgG) with an illness consistent with a clinical description of known LF cases. Some cases were also recorded from Medecins Sans Frontieres (MSF), Hanga town, Kenema District. We defined a person as a control who lived within a one-kilometre radius of the case household and who had not shown any symptoms compatible with LASV infection in the past 3 weeks.

Between June 2021 and January 2022, we enrolled cases and controls from Kenema districts (**Fig 1**). After reaching the case’s house, we explained the objective of our study and requested a signed consent. If the case had died, we collected the data from the closest person related to the deceased person during their illness. In most cases, the closest person was one of the parents or siblings.

**Fig 1:**
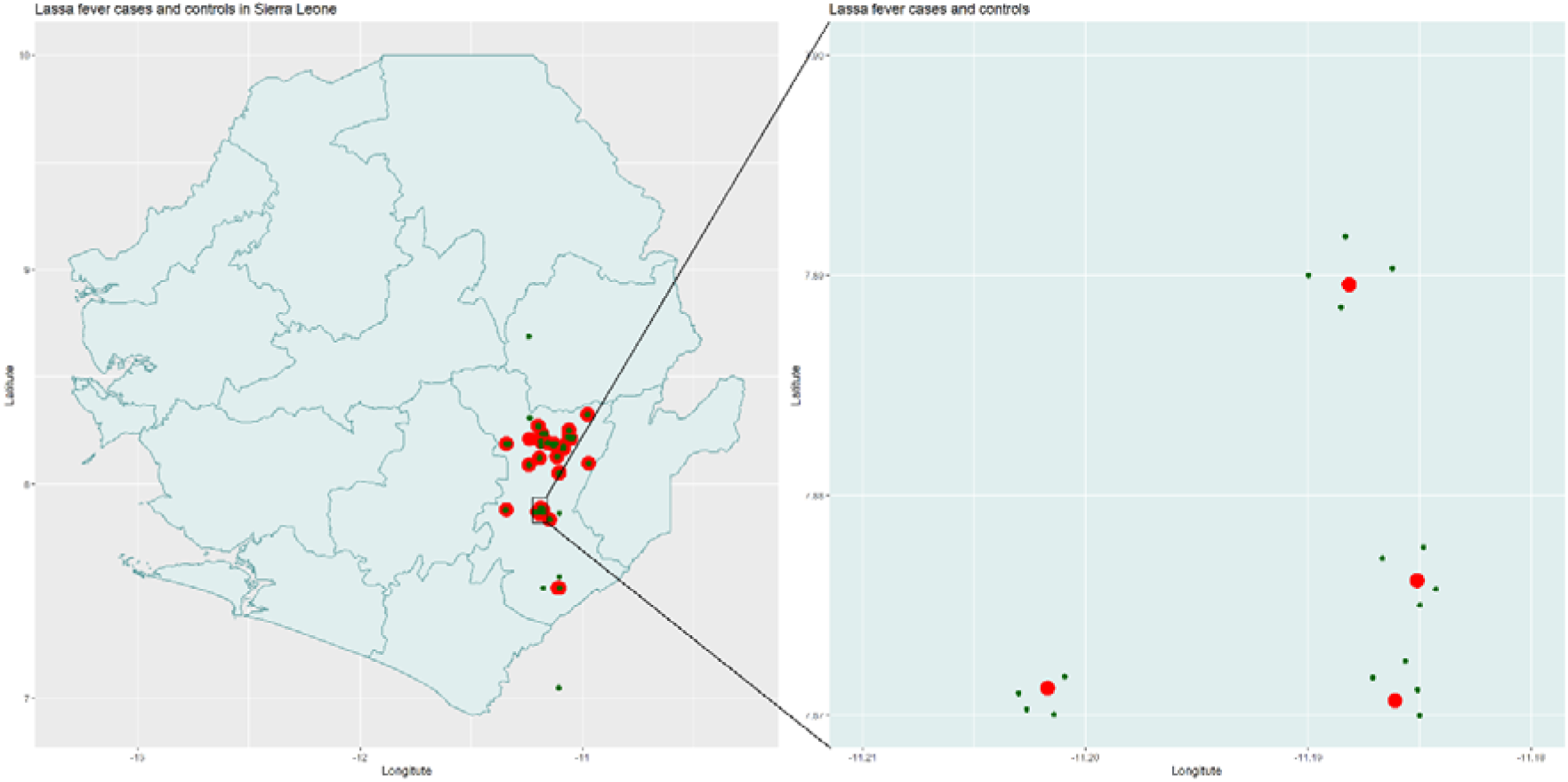
Map of Sierra Leone showing the location of Lassa fever virus infected and their health controls in Kenema District. For each case patient four controls were enrolled within one kilometre of the case household.

After obtaining written informed consent, we interviewed the cases or the closest person of the case using a structured questionnaire containing 51 questions and 11 of the questions had multiple sub-questions. The team inquired about the demographic information of the case (age, sex) and their exposure history in 3-weeks days before the onset of illness including the presence of rodents (rats or mice) in their households, rodents’ activities, having animal contact, presence of cats and dogs at households, involvement with bushmeat (hunting, processing or eating), palm juice processing and the physical location of the household including the estimated number of palm trees around 500m radius of the cases house. We recorded the location of the case house by obtaining their coordinates using handheld global positioning system devices. Cases (or the closest person of the deceased case) who disagreed to provide an interview were not included in this study.

For each case, the team enrolled four individuals as controls from a 1 KM radius of the case’s location. We walk in each of the four directions from the case house (North South, East and West). From each direction, we enrolled one control randomly. We defined a control as a person living within a 1 KM radius area from the identified case house and who did not show any symptoms compatible with LASV infection in the past 3 months. After the agreement and signing of the written informed consent, we administered the same questionnaire used for the case. In one instance, two cases were enrolled from the same household, and we enrolled only 4 controls from them.

Individuals were excluded as controls if they had tested positive for LASV-specific antibodies (IgG or IgM) or PCR in their lifetime or had clinical signs/symptoms compatible with LF infection including fever, malaise, headache, sore throat and muscle pain, vomiting, nausea and diarrhoea, and haemorrhage in 3 weeks before and after the LF case was identified. In case, the approached control was not enrolled, we walked in the same direction to identify another control.

## Variables of interest

1. ***Exposure to rodents:*** *Mystomys natalensis* mice are known reservoirs of LASV. We hypothesised that the presence of rodents and increased interaction with rodents will increase the risk of LASV infection. During our pre-testing of the questionnaire, we identified that people can not differentiate between rats and mice, and for that reason we used local language and description of each species to understand the exposure to mice and rats. Collectively, we had eight questions regarding exposure to rodents and rodents’ activity in their household including the presence of rodents (either rats or mice), frequency of rodents observed, and contact with rodents (touched, eaten, or processed).
2. ***Exposure to animals:*** We were interested to understand whether contact with other animals might be associated with LASV infection and thus included questions on exposure to peri-domestic and domestic animals including monkeys, dogs, squirrels, bats, sheep, goats, cattle, and chicken.
3. ***Bushmeat:*** Bushmeat has been considered as a practice associated with spillover of several zoonotic pathogens. We asked whether individuals were involved in hunting wild animals, processing wild animal meat, and the business of wild animals or meat.
4. ***Infected human:*** We hypothesized that contracting a LASV-infected individual would increase the risk of LASV infection and thus asked whether the subjects were exposed to LASV-confirmed cases 21 days before the onset of illness of the case individual.
5. ***Palm tree and palm juice:*** Palm tree or juice are not known to be associated with LASV infection. However, the presence of palm trees around the household indicates presence of rodents around the households. Also, rodents, especially squirrels or occasionally mice can contaminate the palm juice collecting pot. Thus, we hypothesized that people involved with palm juice collection, processing and business are at increased risk of getting LASV infection.
6. **Demography:** A large proportion of LF cases are mild and asymptomatic [22] and lifetime cumulative exposure to LASV might act as a protective factor for the older population. We hypothesized that being younger in age and female increases the risk of LASV infection [22].
7. **Presence of Cat(s) in the household:** Cats are reared to control rodents in households. We hypothesized that having a cat in the household would reduce burden of rodents in the household and thus contribute as reducing the risk of LASV infection.
8. We have dropped a variable from the final multivariate logistic regression model if the variable.

a. had less than 10% response
b. had temporal embigiuity and
c. was not biologically plausible

## Data analysis

We reported numbers and percentages for categorical variables. For continuous variables, we used to mean with inter-quartile range (IQR). We performed a univariable analysis of variables for reporting the odds ratios (ORs) and the 95% confidence interval (CI) using logistic regression. To build the final regression model, we developed a hypothetical causal diagram by including the variables that are biologically plausible to cause LASV infection (**Fig 2**). We included all eight biologically plausible variables in the conditional multiple logistic regression model irrespective of its significance in univariate analysis to estimate adjusted matched odds ratios and 95% CI. We included only one rodent exposure-related variable (presence of rodent-related exposure in the household) in the final model as other variables indicating the degree of exposure to the households (e.g., Frequency of observing rats and mice (1-2 times vs more than) or rodent activity at the house (observed rat holes, nest, droppings, pups and food damage by rodents). None of the comorbidities [diabetes, hypertension, arthritis] was eligible for inclusion in the model (with more than 50% missing responses). The data analysis was performed in the statistical software STATA version 17. Conditional logistic regression analysis was conducted using ‘clogit’ function by including all controls of each case as group variables.

**Fig 2:**
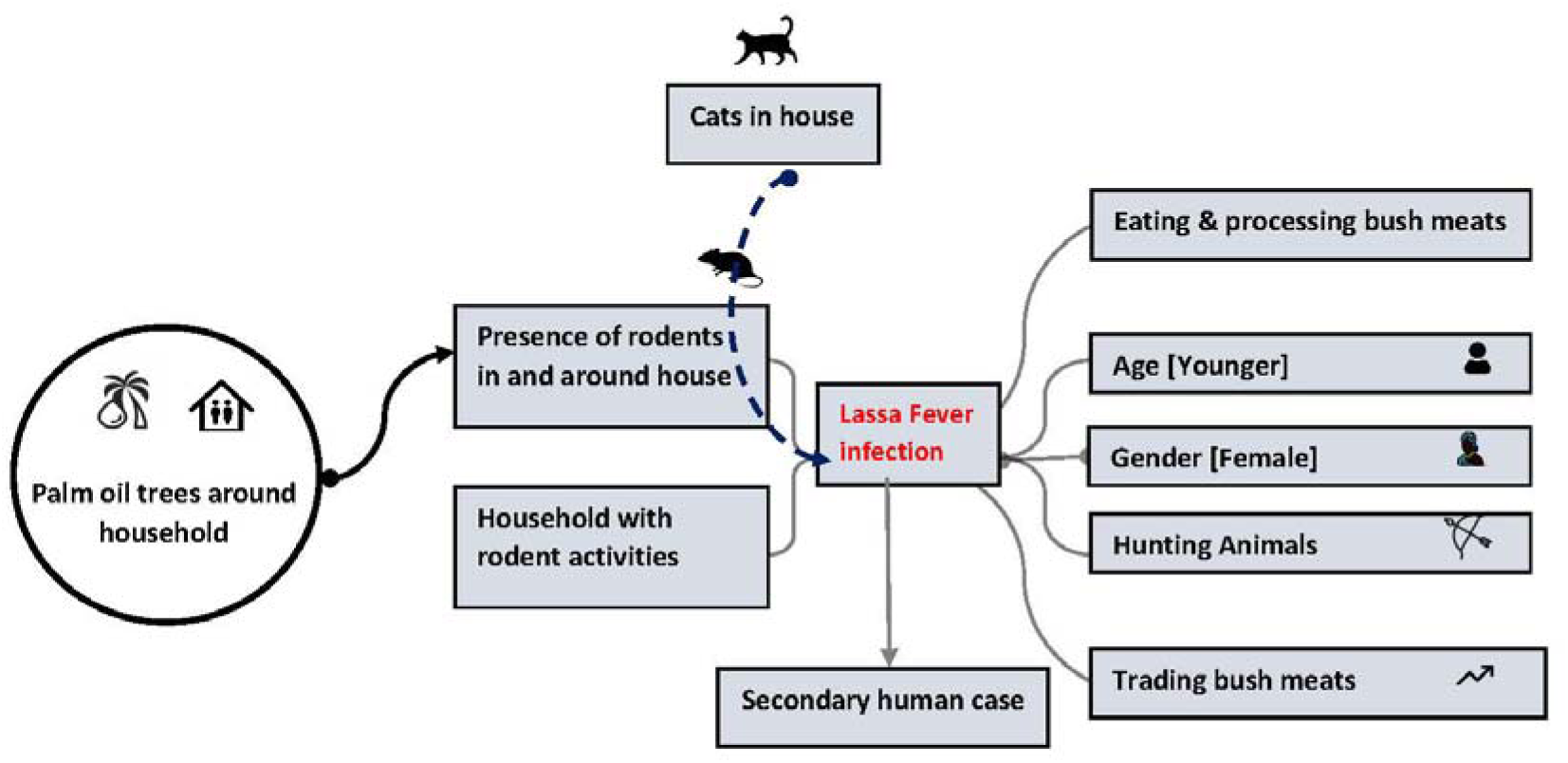
Hypothetical causal relationship between different biological and environmental factors (variables) and clinical Lassa fever infection in Sierra Leone. A solid line indicates a direct relationship between variables. For example, a higher number of palm oil trees is probably associated with the presence of a higher number of rodents in the neighbourhood which ultimately results in the presence of rodents in households. The dotted line indicates interference with other variables. For example, the presence of cats in the house could control the number of rodents in the households and thus could reduce the risk of Lassa virus infection.

### Ethical approval

This study was approved by Sierra Leone Ethics and Scientific Review Committee on 31^st^ October 2019 and the Clinical Research and Ethical Review Board of the Royal Veterinary College, University of London, United Kingdom on 27^th^ March 2022 (URN 2019 1949-3).

## Results

We enrolled 37 cases and 140 eligible controls. Of the 37 cases 23 died of LASV infection, indicating a case-fatality ratio of 62%. The mean age of the deceased cases was 17.0 (interquartile range [IQR]: 3.3-24.0) years, while the mean age of the survivors was 21.1 (IQR: 11.5 - 28.0) years. Of the 37 cases, 36 were hospitalized, 33 had fever, 28 had body aches, 21 had joint pain, 11 had vomiting, 10 had coughing and 4 had bleeding from natural orifice. The patient stayed on average 11.6 (IQR: 7-14) days in hospital before death or discharge. On average, the survivor had to stay 12 days (7.0-13.5) in the hospital whereas the deceased stayed 8.7 days (5.5-9.2) before death. None of the cases or controls had visited another confirmed LASV case or visited any hospital 21 days before the onset of illness of the case patient. Except for one control respondent, all participants have heard of the name Lassa fever.

More than 64% (n=24) of the cases and 52% (n=74) of the controls were female. Compared to the controls, the cases were younger in age (28.8 vs 19.4 years, p=0.01). Cases reported the presence of rodents (combined rats and mice) more frequently than the control in the household in past 3 weeks (83% vs 47%, p<0.01). Case also observed a higher frequency of daily observation of rodents in the household (40.7% vs 72.9%, p<0.01) **(Table 1)**. Cases and controls did not differ in terms of exposure to wild meats including hunting, processing, eating, and/or trading (24.2% vs 18.9%, p=0.63), or having a cat in the household (48.6 % vs 55.7%) **(Table 1)**. We also explored the relationship between several exposure variables including households with cats and reporting rodent activities. Of the 96 households that reported having a cat, only 38% (n=38) observed rodents’ activities in their household compared to 73% (n=58) without any cat in the household (p<0.001).

**Table 1:** Demographics and other important variables of Lassa fever infected cases vs. control individuals in the Kenema district of Sierra Leone identified from January 2018 to December 2021.

|  | Variables | Cases (%)<br>(N=37) | Controls<br>(%)<br>(N=140) | P-value | Matched Odds<br>ratio (95% CI) |
| --- | --- | --- | --- | --- | --- |
| 1 | Age of subject in years, mean (standard deviation) | 19.5 (±18.8) | 28.9 (±20.8) | <0.01 | 0.993 (0.989-0.996) |
| 2 | Female gender (%) | 24(64.8%) | 51 (52.8%) | 0.29 | 1.5 (0.71-3.2) |
| 3 | Presence of rodents (rats or mice) in the household in the past 3 weeks | 31 (83.8%) | 67 (47.8%) | **<0.001** | **6.8 (2.5-18.6)** |
| 4 | Frequency of observing rats and mice (1-2 times vs more than twice daily) | 27 (72.9%) | 57 (40.7%) | **<0.001** | **3.9 (1.81-9.12)** |
| 5 | Rodent activity at house (observed rat holes, nest, droppings, pups, and food damage by rodents) | 23 (57.5%) | 57 (40.7%) | **0.01** | **2.6 (1.2-5.9)** |
| 6 | Having a domestic animal contact (processing, killing, or cooking animals) in the past 3 weeks | 25 (67.6%) | 86 (61.4%) | 0.60 | 1.60 (0.61-4.20) |
| 7 | Touching of wild or peri domestic animals' animals (mice, rats, monkeys, squirrels, or other wild animals) in the past 3 weeks | 7 (18.9%) | 11 (7.8%) | 0.09 | 2.7 (0.84-9.9) |
| 8 | Presence of a cat in the household | 18 (48.6%) | 78 (55.7%) | 0.56 | 0.75 (0.36-1.55) |
| 9 | The mean number of palm oil trees around 100 m radius of the household | 9.75 | 4.10 | 0.36 | 1.03 (0.98-1.13) |
| 10 | Exposure to bush meats (Hunting, eating, processing, and trading bush meats) | 7 (18.9%) | 34 (24.2%) | 0.63 | 0.92 (0.29-2.91) |
| 11 | Any member of your family collected palm oil juice | 15 (37%) | 38 (27.1%) | 0.16 | 1.83 (0.84-3.87) |
| 12 | Presence of the dog in the household | 5 (13.5%) | 21 (15.0) |  | 1.06 (0.36 -3.12) |

The multivariable analyses provided evidence of an association between odds of LASV infection and the presence of rodents in the household (mAOR: 11.1 (95% CI: 2.8-42.4) and age in years (mAOR: 0.99 (95%: 0.98-0.99) **(Table 2)**. Other variables, including gender, showed no evidence of association with odds of infection following adjustment for other variables **(Table 2)**.

**Table 2:**
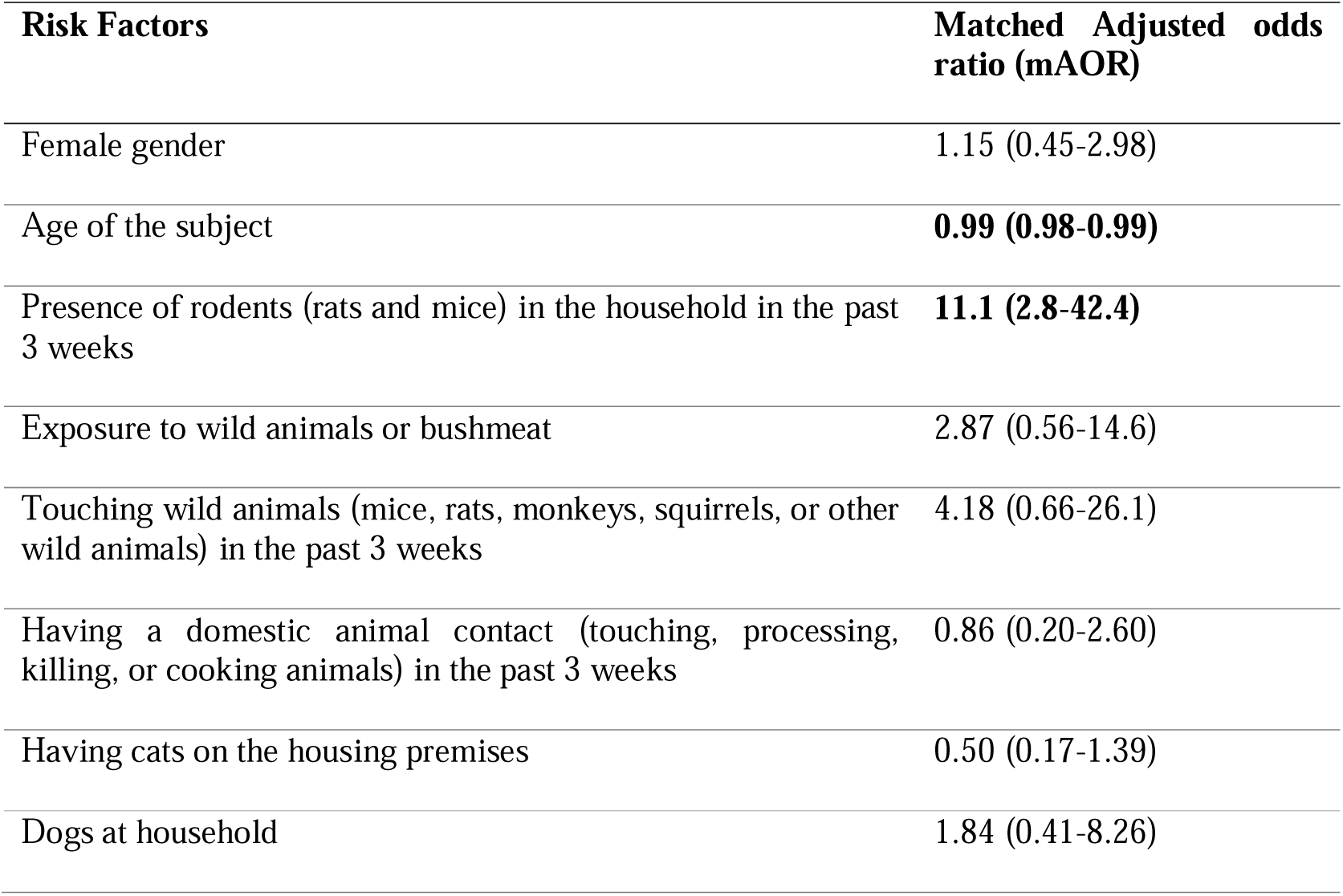
The factors associated with Lassa fever infection in humans in a multiple logistic regression analysis. Cases were reported between Jan 2019 and December 2021 and data were collected between Dec 2021 and August 2021.

## Discussion

We identified rodent access in the household markedly increased (e.g. by 11-time) the risk of LASV infection in rural Sierra Leone. We further found that the younger the individual the higher the risk of developing fatal LASV infection. In the univariable analysis, we found a dose-response of rodent activity (observing rats and mice 1-2 times vs more than twice as well as observing rat holes, nest, droppings, pups, and food damage by rodents) and LASV infection risk, where higher daily observing of rodent activity was associated with a higher risk of LASV infection. This is highly plausible and supports our current understanding of LASV transmission in rural West African settings.

Lassa fever virus has been circulating in West Africa for the last six decades or possibly hundreds of years posing a continuous public health threat to the region. However, the identification of risk factors for LASV infection using a systemic case-control study is very rare. One possible obstacle for such a study is that LASV infection, when clinically manifested, is very severe and often fatal [22], and collecting data from the cases is challenging. Another potential barrier is that a vast majority of the cases are asymptomatic [22] and thus enrolment of cases is problematic with a potential of subject misclassification bias without laboratory confirmation. Nonetheless, a case-control approach has been proven ideal where knowledge regarding potential risk factors remains limited and the study of a broad range of risk factors linked to different causal pathways is warranted. Our study, despite some of these existing limitations, attempted to identify risk factors for LASV infection and helped in generating several hypotheses which need further systematic studies.

The multimammate mouse, *Mastomys natalensis* has been considered as the key reservoir of LASV, with humans being infected directly or indirectly through fluids of the mouse such as urine, saliva, and blood [23]. In the same study areas in Sierra Leone, 92% of dwellers reported the presence of rodent species inside the households and 57% of the trapped rodent species were *M. natalensis* [23]. A recent rodent trapping study in the same areas identified 2.8% of trapped *M. natensis* were positive for LASV [24] underscoring a high chance of rodent-human transmission.

Several cross-sectional studies established the link between exposure to rodents and LASV infection. A study conducted in rural Guinea in the 1990s identified hunting peri-domestic rodents and consumption of rodents as potential risk factors [25]. Another study further identified household-level risk factors for increased abundance of rodents, including households having more than 8 holes and the presence of rodent burrows [26]. Thus, our findings support the current understanding of household-level rodent-human transmission. In our enrolled study population, none of the cases reported visiting a hospital or sick people 21 days before onset of illness indicating a primary spill-over of the LASV infection.

We found age as a protective factor of LASV infection (mAOR: 0.99), with an additional year of age the odds of LASV infection decreased by 1%. Further, the deceased cases were younger than the survivors (17.0 years vs 21.1 years). A large proportion of LASV infections are asymptomatic [22] and thus older aged people possibly acquire immunity against LASV infection through life-time cumulative exposure to the virus.

Although the final multivariable analysis did not provide evidence of other variables being associated, our study raised several potential hypotheses. For example, cats have been promoted in rodent killing programs in West Africa but whether the cats can reduce the burden of rodents, or it become infected itself and be a source of transmission has not been studied. In our univariate analysis, we found that households having a cat reported lower presence of rodents’ activity in the premises (73% vs 38%, p<0.05) and contributed in a lower rate of LASV infection (48.6% vs 55.7%) although not statistically significant (p=0.56). However, this could be an economic artifact because the presence of a cat in the household indicates their economic strength which might influence better housing that prevents access to rodents. Ideally, this association between exposure variables is considered to be a confounder. However, we kept both variables (rodents and cats) in the final regression model as both variables could affect LASV exposure. This will be interesting to study further details where the presence of cats (or a number of cats) in the households reduces household-level rodent infestation sufficient to control LASV infection. Our study further showed that LASV-case households had a higher number of palm oil trees around the 500m radius area of their household. Although palm tree itself is not a risk factor, increased palm tree possibly facilitates a higher number of rodents nestling in the bushes. Possible contamination of juice collected from oil palm tree should be studied for evidence of LASV.

Our study did not detect any increased risk for exposure to bush meat, the presence of dogs in the households, and family members’ involvement with palm oil juice preparation or business. However, the lack of evidence in our study does not necessarily rule out these variables as potential risk factors for LASV infection in other settings or another well-designed study conducted in the same settings, as some of these variables were identified as a risk factor in other country and the statistical power of our study was limited given the small sample [25].

This study has several limitations. First, we did not confirm the controls as test negative. This is critical when we know that a large proportion of LASV infections are asymptomatic and people living in the endemic areas like Kenema district might have a high prevalence of LASV exposure (e.g.20.1%) [27]. We tried to minimize potential classification bias by asking for all the clinical signs compatible with Lassa fever infection. As LASV is a serious concern in the community, we believe people take attention to their illness when a case of LASV is identified in the community. All our controls were enrolled from the same community, within a 500m radius of the case individual. However, our study could not adjust the possible misclassification associated with asymptomatic infection among controls. Second, like all other case-control studies, our study might have included recall bias. To avoid recall bias, we physically verified some of the questions. For example, access to rodents in the households was observed and questions were placed in a way that the respondent could self-verify his response. Thus, we believe recall bias was minimal in our study findings. Finally, we took verbal autopsies of the cases who died of LASV infection which might lead to some information bias. However, most questions we included were answerable by any nearest individuals as most LASV exposure is household level (e.g., rodents’ access to household) or through group exposure (e.g. bush meat).

## Conclusion

Rodent access to households is likely a key risk factor for LASV infection in rural Sierra Leone and potentially in other countries within the region. Younger people are at high risk of developing symptomatic LF infection. Controlling the rodent population in rural areas endemic to LASV emerges as a critical measure to prevent household-level exposure of humans to LASV. Vaccines when available should target the younger aged population as a priority. We recommend studying the role of cats in the prevention of rodents thereby reducing the overall risk of LASV infection in endemic countries.

## Data Availability

All data produced in the present study are available upon reasonable request to the authors

## Acknowledgment

This study was funded by the European and Developing Countries Clinical Trials Partnership (EDCTP2) programme, which is supported under Horizon 2020, the European Union’s Framework Programme for Research and Innovation through PANDORA-ID-NET Consortium (EDCTP Reg/Grant RIA2016E-1609). All authors are member of PANDORA-ID-NET. NH, RA, AYO and JG are members of the International Development Research Centre, Canada’s grant on West African One Health Actions for understanding, preventing, and mitigating outbreaks (109810-001). AZ is a National Institutes of Health Research senior investigator.

## Author contribution statement

NH, RK, and RA originally planned the study, and DS collected, and created an Excel version of the field data. NH and JG analyzed the data. NH and DS prepared the first draft manuscript, and all co-authors reviewed the draft manuscript. All authors approved the submission of the manuscript.

## Ethical approval

This study was approved by the Ethical Committee of Sierra Leone Ethics and Scientific Review Committee on 31^st^ October 2019 and the Clinical Research and Ethical Review Board of the Royal Veterinary College, University of London, United Kingdom on 27^th^ March 2022 (URN 2019 1949-3).

